# Multiomic Analysis Identifies a High-Risk Metabolic and TME Depleted Signature that Predicts Early Clinical Failure in DLBCL

**DOI:** 10.1101/2023.06.07.23290748

**Authors:** Kerstin Wenzl, Matt Stokes, Joseph P. Novak, Allison M. Bock, Sana Khan, Melissa A. Hopper, Jordan E. Krull, Abigail R. Dropik, Janek S. Walker, Vivekananda Sarangi, Raphael Mwangi, Maria Ortiz, Nicholas Stong, C. Chris Huang, Matthew J. Maurer, Lisa Rimsza, Brian K. Link, Susan L. Slager, Yan Asmann, Patrizia Mondello, Ryan Morin, Stephen M Ansell, Thomas M. Habermann, Andrew L. Feldman, Rebecca L. King, Grzegorz Nowakowski, James R. Cerhan, Anita K. Gandhi, Anne J. Novak

## Abstract

**PURPOSE:** 60-70% of newly diagnosed diffuse large B-cell lymphoma (DLBCL) patients avoid events within 24 months of diagnosis (EFS24) and the remainder have poor outcomes. Recent genetic and molecular classification of DLBCL has advanced our knowledge of disease biology, yet were not designed to predict early events and guide anticipatory selection of novel therapies. To address this unmet need, we used an integrative multiomic approach to identify a signature at diagnosis that will identify DLBCL at high risk of early clinical failure.

**PATIENTS AND METHODS:** Tumor biopsies from 444 newly diagnosed DLBCL were analyzed by WES and RNAseq. A combination of weighted gene correlation network analysis and differential gene expression analysis followed by integration with clinical and genomic data was used to identify a multiomic signature associated with high risk of early clinical failure.

**RESULTS:** Current DLBCL classifiers are unable to discriminate cases who fail EFS24. We identified a high risk RNA signature that had a hazard ratio (HR, 18.46 [95% CI 6.51-52.31] *P* < .001) in a univariate model, which did not attenuate after adjustment for age, IPI and COO (HR, 20.8 [95% CI, 7.14-61.09] *P* < .001). Further analysis revealed the signature was associated with metabolic reprogramming and a depleted immune microenvironment. Finally, WES data was integrated into the signature and we found that inclusion of *ARID1A* mutations resulted in identification of 45% of cases with an early clinical failure which was validated in external DLBCL cohorts.

**CONCLUSION:** This novel and integrative approach is the first to identify a signature at diagnosis that will identify DLBCL at high risk for early clinical failure and may have significant implications for design of therapeutic options.

## INTRODUCTION

While the majority of DLBCL patients are potentially cured after standard therapy, there remains a subset of patients who do not respond to front line treatment^1^. The approximate 70% of DLBCL patients who avoid retreatment, progression, relapse, or death within 24 months of diagnosis (termed event-free survival at 24 months or EFS24) have a good prognosis while the remaining 30% have a very poor outcome.^2^ Using clinical factors we developed and validated the International Prognostic Index for EFS24 (IPI24),^3^ which can be used at diagnosis for personalized risk prediction.^3^ Beyond clinical characteristics,^4^ molecular features associated with DLBCL prognosis include cell-of-origin (COO)^5, 6^ and *MYC, BCL2,*or *BCL6* translocation status, or *MYC* “double hits (DH)”,^7–9^ with DH-DLBCL now considered as a distinct entity, High Grade B Cell Lymphoma (HGBCL).^10^ More recent molecular classification of DLBCL based on genomics, expression profiles, and tumor microenvironment has further refined our understanding of DLBCL heterogeneity and biologic underpinnings.^11–18^ While these studies have advanced our understanding of DLBCL, none were designed to identify early failures (EFS24) after frontline standard of care therapy, which is of great interest for patient management and could provide biologic insight and identification of therapeutic targets. In addition, optimal utilization of novel treatment strategies that are somewhat agnostic to tumor biology, such as chimeric-antigen receptor T cell (CAR T) or bispecific T-cell engager (BiTE) antibody therapy, could benefit from identification of patients at high risk of early failure.

To identify a biologic signature of early clinical failure, we used next-generation sequencing (NGS) data generated on newly diagnosed (ndDLBCL) and relapsed/refractory DLBCL (rrDLBCL) tumors, combined with integrative computational approaches. Based on this signature, patients at diagnosis can be categorized into low, intermediate, or high-risk groups for early clinical failure (EFS24) and inferior overall event-free (EFS) and overall (OS) survival, independent of IPI, COO, and other known factors.

## METHODS

### Study Populations

The overall study design is shown in Figure S1. We used clinical and NGS data from diagnostic tumors from 444 DLBCL patients from the University of Iowa and Mayo Clinic Lymphoma Specialized Program of Research Excellence (SPORE) Molecular Epidemiology Resource^19^ (MER, n= 433) or from NCT00670358 (n=11)^20^, herein referred to as MER. Patients provided written consent at study enrollment, including use of clinical samples. Individual patient level data is shown in Table S1, all identifiers are coded, and methods are in Supplemental Methods. We also used NGS data from tumor samples at the time of relapse (rrDLBCL, any line of treatment), consented to the MER (n=61), banked in the Mayo Lymphoma Biobank (waiver of consent) (n=50), or consented to the CC-122-ST-001 clinical trial (n=32, NCT01421524). NdDLBCL validation cohorts include those from the BCCA (EGAS00001002936), Duke (EGAD00001003600), and REMoDL-B (GSE117556).^18, 21, 22^

### DNA Sequencing and Analysis

For WES, we used paired tumor (FFPE) and germline (extracted from peripheral blood) DNA; sequencing was conducted at Expression Analysis, Inc (Durham, HC, USA), as described in Supplemental Methods. After quality control, WES data on 341 ndDLBCL was included. We also used existing WES data generated at Mayo on 19 additional ndDLBCL tumors, as well as previously analyzed WES data on Mayo DLBCL cases from Lohr et al (n=16) and Hartert et al (n=28).^23, 24^ The final analysis cohort included data from 404 ndDLBCL. Genes included for analysis are shown in Table S2. Mutation calls from the BCCA (n=121) and Duke (n=441) cohorts were provided by Dr. Morin, mutation calls from REMoDL-B (n=400) were obtained from Sha et al.^22^ Copy number analysis (CNA) was carried out using the Nexus Copy Number (Biodiscovery) software, detailed in Supplemental Methods.^24–26^ Classification methods for LymphGen and HMRN and tools used for analysis are in Supplemental Methods.

### RNA Sequencing and Analysis

RNA was extracted from FFPE tissue sections and sequencing was performed at Expression Analysis, Inc as described in Supplemental Methods. Sample and RNA QC is shown in Figure S2 for a final cohort of 321 ndDLBCL and 143 rrDLBCL cases. For validation, we used data from BCCA provided by Dr. Morin (n=121); Duke, downloaded from EGA and processed in the Mayo Clinic Bioinformatics Core (n=442); and REMoDL-B, downloaded and processed on the NCBI GEO website (n=928). ^18, 21, 22^ Data analyses, including the weighted gene correlation network analysis (WGCNA) and differential gene expression analysis, are described in Supplemental Methods.

### Statistical Analysis

EFS was defined as time from diagnosis to disease relapse/progression, retreatment, or death, and EFS24 was defined as EFS status at 24 months after diagnosis.^2^ OS was defined as the time from diagnosis to death from any cause. EFS and OS were evaluated using Cox proportional hazards models and Kaplan Meier curves. EFS24 associations survival curves were truncated at 24 months. Differences in survival curves were evaluated using the log-rank test. For enrichment analysis of categorical variables, the Chi Square or Fisher’s exact test was used. Comparative analyses were carried out using either the Wilcoxon or Kruskal-Wallis test. A *P*-value <0.05 was considered statistically significant unless otherwise stated. Analysis were performed using R and GraphPad Prism.^28^

## RESULTS

### Cohort Description and Performance of Published Molecular Classifiers for Early Clinical Failure

A summary of available molecular and genetic features on the 444 ndDLBCL is shown in Figure S3A. The median age at diagnosis was 64.5 years, 57% were male, and all were treated with immunochemotherapy; full clinical details are summarized in Figure S3B. During a median follow-up time of 82.8 months (for living patients), 168 (37.8%) had an event and 112 (25.2%) failed to achieve EFS24 (early clinical failures). While some DLBCL molecular classifiers have been associated with prognosis, none were designed to discriminate early clinical failures. In our cohort, COO^5^ (overall, *P* = .05, compared to GCB, ABC HR, 1.63 [95% CI 1.08-2.45] *P* = .019, and Unclassified HR, 1.50 [95% CI 0.84-2.67] *P* = .169), DH FISH (DH FISH+ HR, 1.93 [95% CI 0.96-3.86] *P* =.064), and DH Sig+^29^ (HR, 1.58 [95% CI 0.93-2.68] *P* =.093) showed nominal associations with EFS24 in expected directions (Figure S4). The recently developed DLBCL molecular classifiers HMRN, LymphGen, and EcoTyper B Cell State showed expected distributions (Figure 1 A-C top panel), but were not associated with EFS24 (Figure 1 A-C, lower panel) and EFS24 failures spread across groups within each classifier as shown in Sanky plots (Figure 1 A-C, middle panel top). The genomic classification based on Chapuy et al was only available on 41 cases and therefore was not included for further analyzed.^12^

**Figure 1.**
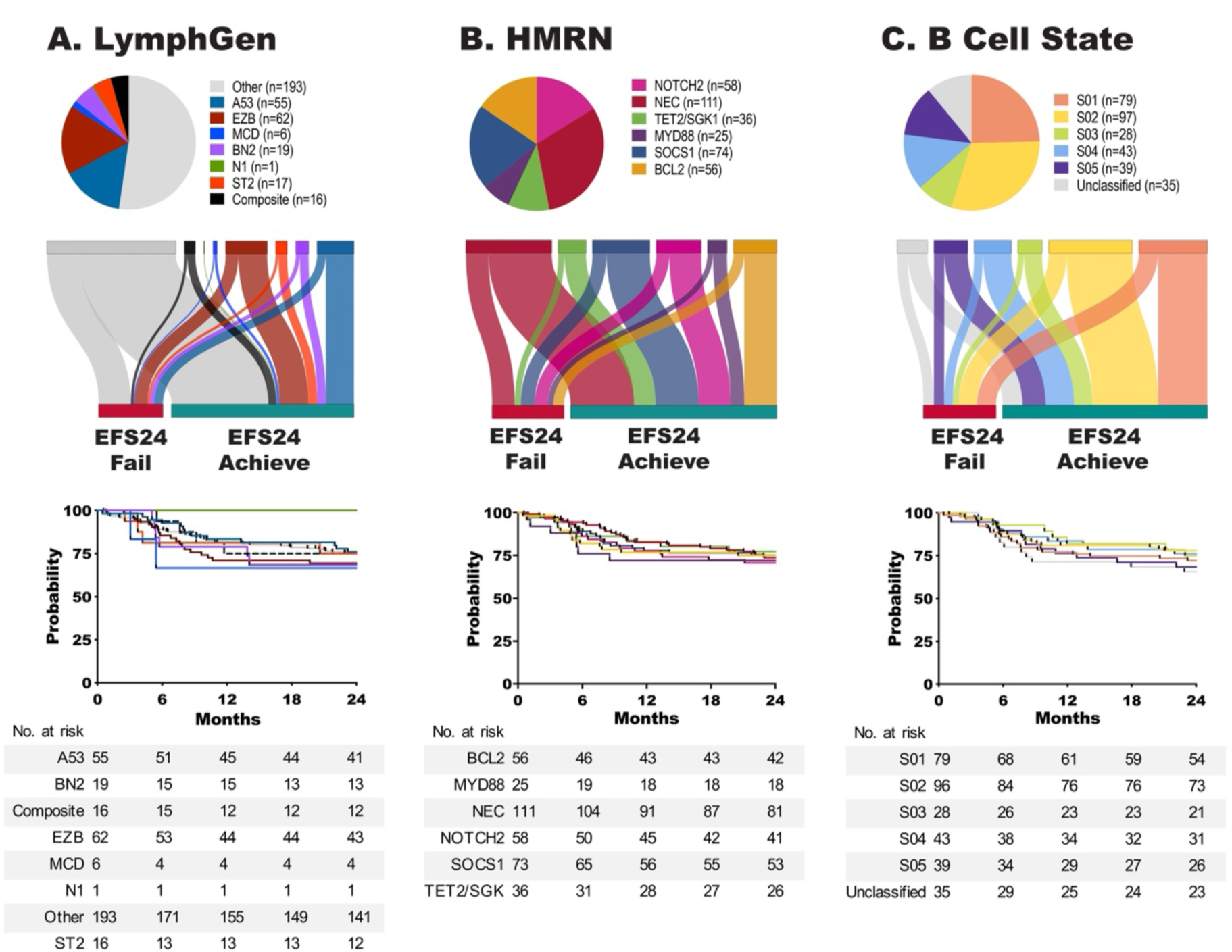
Current DLBCL Classifiers Do Not Discriminate Early Clinical Failures. LymphGen (A), HMRN (B), and EcoTyper B Cell State (C) classification of ndDLBCL. Pie charts (upper panel) show distribution of cases for each classifier. Sankey plots (middle panel) show the distribution of EFS24 fail or achieve cases for each classifier. Kaplan Meier analysis (lower panel) of EFS24 for each classifier, LymphGen *P* = .96, HMRN *P* = .98, and B Cell State *P* = .73

### WGCNA Analysis of DLBCL

Because the existing molecular classifiers were at best weak predictors of EFS24, we analyzed the RNA-seq data from our ndDLBCL cohort (n=321) using WGCNA,^30^ a method for identifying biologic networks, or gene modules, by using pairwise correlations between variables (Figure 2A). Unsupervised hierarchical clustering, followed by branch cutting, identified 15 modules with a range of 17 to 2685 genes (Figure 2B). EFS24 as well as COO and DH characteristics were correlated with individual gene modules (Figure 2C). The strongest positive correlations were observed for the cyan module with ABC and the pink module with GCB. The pink module also showed strong correlation with both DH FISH and DH Signature. Genes in the pink and cyan modules are in Table S3. As proof of principle for the utility of WGCNA, we calculated the eigengene gene score for each patient for the two COO-associated modules and found that the scores correlated well with their COO (Figure S5).

**Figure 2.**
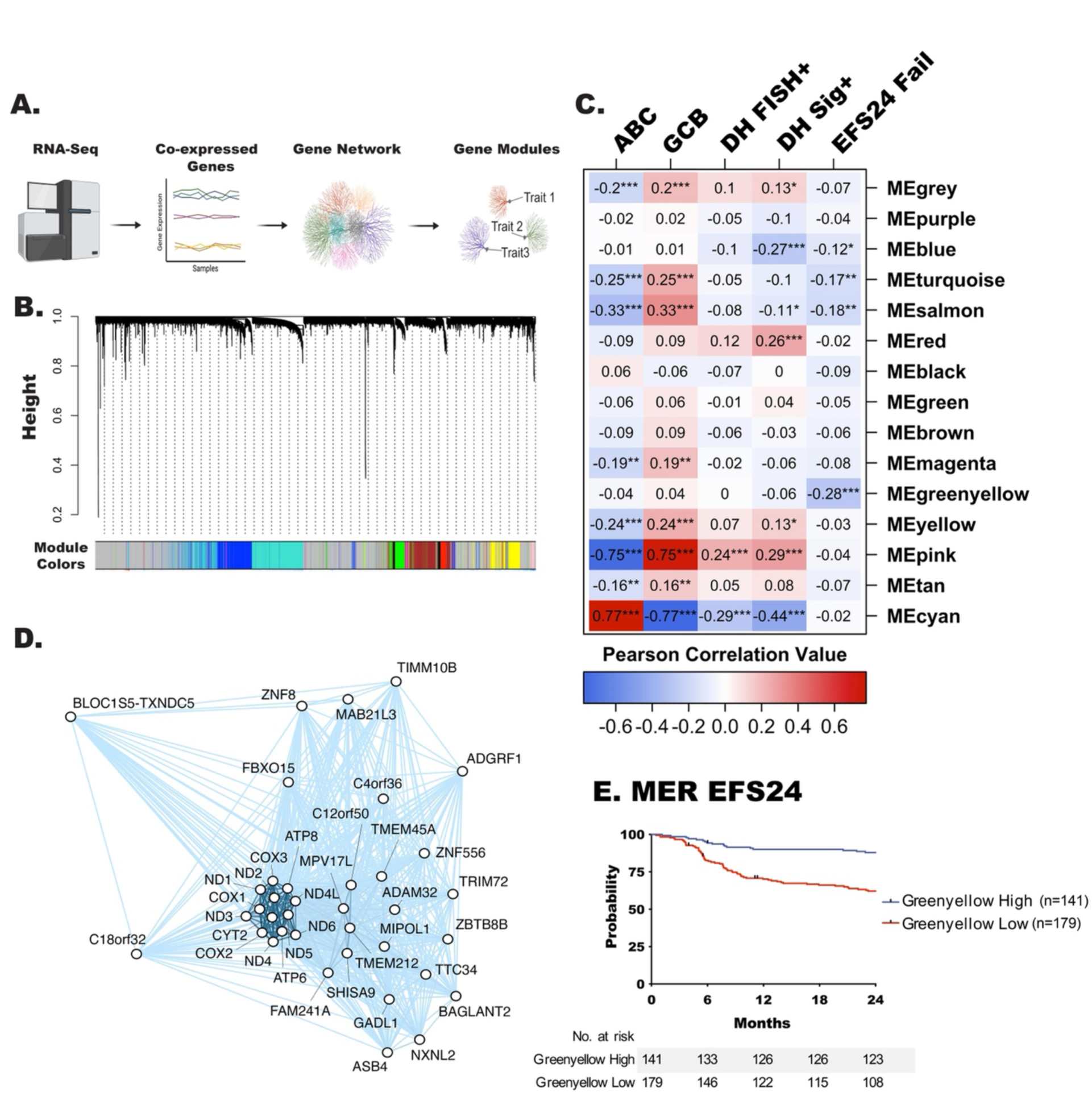
WGCNA Analysis Identifies Biological Modules Associated with DLBCL Clinical Traits. A. Schematic representation of WGCNA analysis workflow, created with BioRender.com. B. Cluster dendrogram showing the 15 identified modules defined by color. The grey module consists of genes that could not be assigned to a co-expression module. C. Correlation of individual WGCNA modules with selected traits (COO of ABC or GCB, n=279, DH-FISH n=252, DH-Sig, n=313, and EFS24, n=321) was performed using Pearson correlation, \**P* < .05, \*\**P* < .001 and \*\*\**P* < .0001. D. Correlation network representation of greenyellow module genes (n=37) analyzed using the igraph R package. E. Kaplan Meier analysis of EFS24 for cases classified as the yellowgreen high or low.

The greenyellow module had the strongest correlation with EFS24 failure (r = −0.28), and was selected for further investigation. Genes (n=37; Table S3) in this module had a negative correlation with EFS24 failure, suggesting that downregulation of their expression may be associated with early clinical failure (Figure 2C). Figure 2D shows the correlation network for the 37 genes in the greenyellow module. To identify an optimal cutpoint for the greenyellow module with EFS24, the eigengene value from this module was calculated for each patient and maxrank statistics (R package) was used^27^ to define high vs. low expression. Figure 2E shows the KM curves based on this cutpoint, such that patients with lower expression of the greenyellow signature genes were more likely to fail EFS within 24 months compared to those with higher expression (HR, 3.67 [95%CI 2.15-6.25] *P* < .001).

### Generation of a High Risk EFS24 Failure Gene Signature

As the WGCNA analysis was unsupervised, we conducted complementary analyses that trained on the EFS24 endpoint using RNA-seq data from both nd- and rrDLBCL cases. We selected protein coding genes that were 1) differentially expressed (FDR<0.05, n=779 genes) between cases who failed (n=84) and achieved (n=237) EFS24; and 2) differentially expressed (FDR<0.05, n=3640 genes) between cases who achieved EFS24 (n=235) and rrDLBCL cases (n=143, Figure 3A). Next, genes common to both analyses were intersected and added to the 37 genes from the WGCNA analysis, ultimately defining a 387 gene signature associated with EFS24 failures and relapsed disease (Figure 3B and Table S4). To score each patient for the gene signature, the R tool singscore was used, which generates a totalscore for all 387 up- and downregulated genes (Figure 3C).^31^ Patients who achieved EFS24 had a lower totalscore (median=0.108), while patients who failed EFS24 (median=0.165) or who had rrDLBCL (median=0.168) had significantly higher totalscores. Next, we divided the cases into three groups based on the distribution of their totalscore by using a cut point of +/-one standard deviation (Figure 3D), which shows that EFS24 failures increase with higher scores. A heatmap of the 387 gene signature, herein referred to as the risk signature, for the low and high risk groups is shown in Figure 3E. A heatmap of all cases is shown in Figure S6.

**Figure 3.**
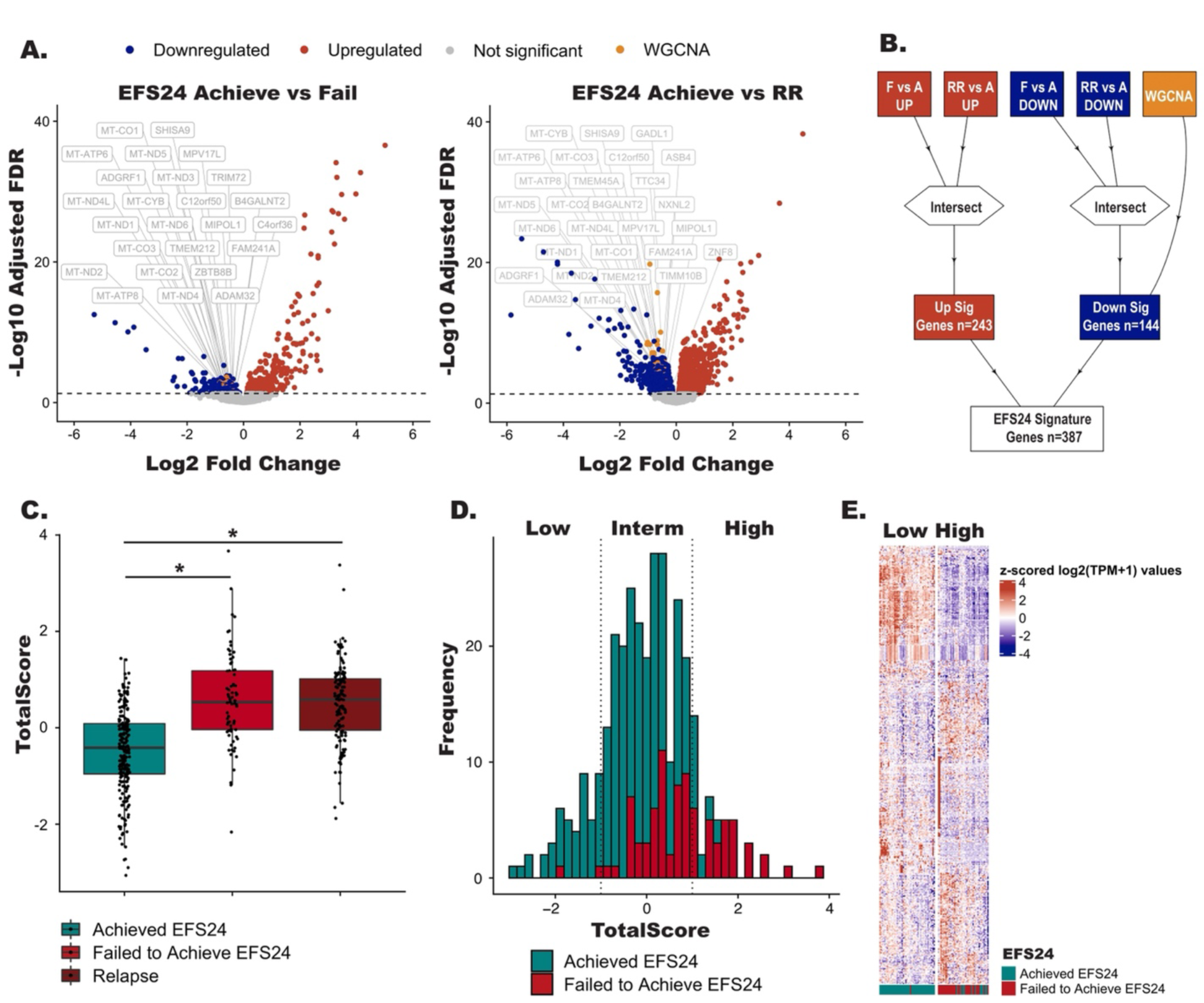
Generation of the RNA Risk Signatures. A. Volcano plots showing differentially expressed genes (FDR <0.05) between EFS24 achieve vs fail or EFS24 achieve vs rrDLBCL cases. Red dots represent upregulated genes and blue dots represent downregulated genes. Grey boxes highlight known lymphoma driver genes. B. Schematic representation of how the 387 gene risk signature was generated. C. Boxplots TotalScores for the risk signature foreach patient. \**P* < .05. D. Distribution of the scaled Totalscores. Vertical lines represent +/-standard deviation which groups the scaled Totalscores samples into high, low and intermediated risk cases. E. Heatmap representing gene expression of RNA signature genes in high k and low Risk samples.

### RNA Risk Signature is Associated with Prognosis

There was a strong association of the RNA signature score with EFS (Figure 4A) and OS (Figure 4B). Compared to low risk, patients with an intermediate (HR, 4.86 [95%CI 1.78-13.24] *P* = .002) or high risk (HR,18.46 [95% CI 6.51-52.31] *P* < .001) signature had inferior EFS, which did not attenuate in the high risk cases after adjustment for COO or IPI (HR, 16.35 [95% CI 5.74-46.56] *P* < .001). Furthermore, results held in analyses stratified on COO, IPI, and after exclusion of HGBCL cases (Figure S7 A-E). The clinical features of cases in each of risk group are shown in Table S5. Next, we attempted to validate our findings using RNA-seq data from cases with available outcome data from BCCA Duke, and REMoDL-B (Figure 4 C-E). Compared to the low risk RNA signature group, patients with a high risk signature in BCCA (PFS, HR, 9.62 [95% CI 2.12-43.55] *P* = .003), Duke (OS, HR, 3.04 [95% CI 1.58-5.83] *P* = .001) and REMoDL-B (PFS, HR, 2.4 [95% CI 1.57-3.68] *P* < .001) had inferior outcomes.

**Figure 4.**
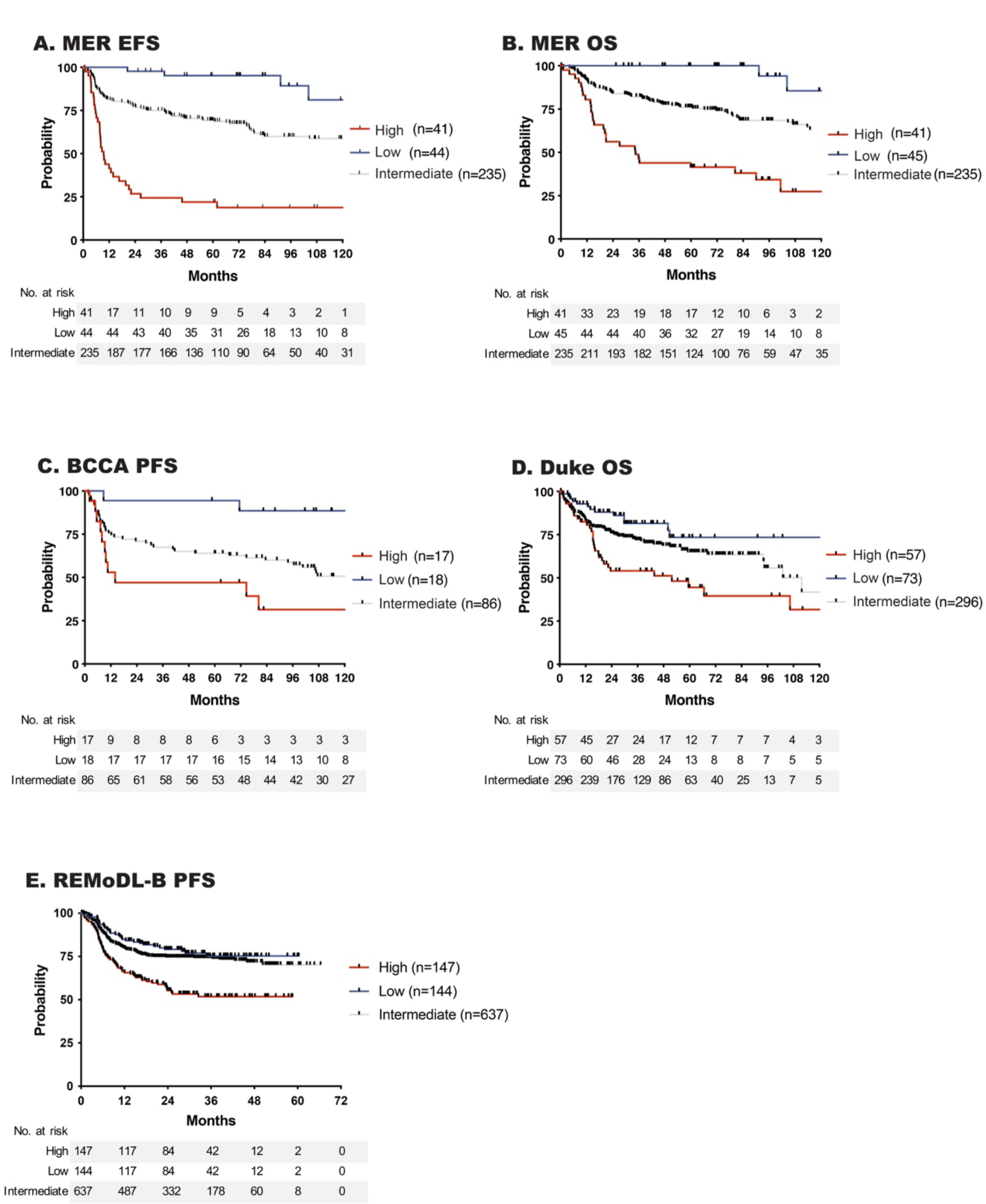
Outcome and Clinical Characteristics of Risk Signature Groups. A. Event free and B. overall survival of MER ndDLBCL cases according to RNA risk signature classification. C-E. Validation of RNA risk signature association with outcome in the BCCA, Duke and REMoDL-B DLBCL cohorts.

### High Risk Cases are Enriched With Metabolic and TME Signatures

To better understand the biology underlying the RNA signature, we conducted additional *in silico* analyses. Based on TME26 scoring, we found a significant enrichment of TME negative cases in the high risk group (Table S5, *P* < .0001), suggesting the importance of overall cellular composition and TME biology in these tumors. To further explore which biological processes drive the high risk signature, we first performed pathway analysis. We first looked at all 3 RNA analyses individually, the WGCNA gene set, and the DEG analyses from Figure 3A. As shown in Figure S8A, oxidative phosphorylation and metabolic processes genes are represented in the greenyellow module. In the DEG analysis between EFS24 achieve vs EFS24 fail we again identified oxidative phosphorylation and metabolic processes (Figure S8B). In the DEG analysis between EFS24 achieve and rrDLBCL, there were genes involved in mismatch repair, NF-κB, and inflammation (Figure S8C). Next, we ran pathway analysis on the 387 genes from our high risk signature and again identified genes that are involved in metabolic processes and oxidative phosphorylation (Figure 5A).

**Figure 5.**
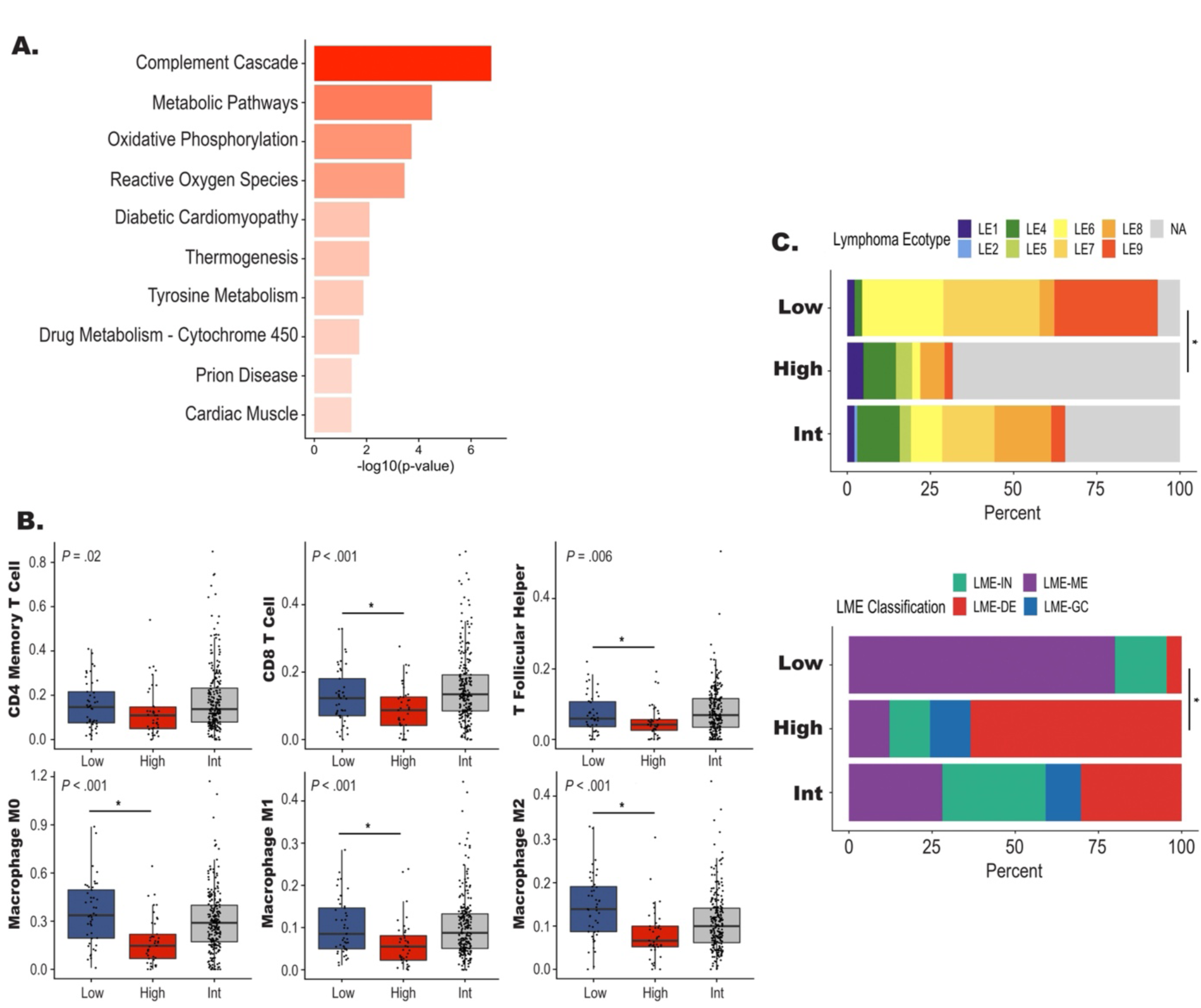
Pathway and TME Characteristics of High Risk Signature DLBCL. A. Bar plot displaying results from overrepresentation analysis for high risk cases. B. Boxplots of individual cell populations identified by CIBERSORTX in the low, high, and intermediate risk groups. *P*-values represent comparison between all three groups performed by a Kruskal–Wallis test and the line represents a \**P* < .05 between high and low risk groups performed by Wilcoxon test. C. Bar plot showing the distribution of Lymphoma EcoType and LME classification in each risk group. The line represents a \**P* < .05 for the comparison of the number of NA or LME-Depleted between the high and low risk groups.

To expand on the TME26 findings and explore the immune composition of the high risk tumors we profiled the TME using CibersortX to assess immune cell content. As shown in Figure 5B, there was a significant decrease in CD4 memory and CD8 T cells, T follicular helper (T_FH_), M0, M1, and M2 macrophages in the high risk group. These results were further supported when we examined the TME using the Lymphoma Ecotype or LME classifiers (Figure S9), both of which identified a significant increase in LME depleted or unclassified TME Ecotype in the high cases vs low risk cases (Figure 5C). Of note, EcoTyper and LME classification were not associated with EFS24 (Figure S9).

### Integration of High Risk Signature With Genetic Features

To determine if our high risk signature was driven by unique genetic features, we used WES and OncoScan data to define their mutation and copy number landscape.

Oncoplots for the mutation and copy number variants are shown in Figure S10A-B. The high risk cases were significantly enriched for mutations in *TP53* and *CREBBP* as well as copy number alterations in 18q21.33 (*BCL2*), 3q28 (*BCL6*), 6q14 (*TMEM30A*), 19q13.42, and 17q24.3 when compared to low risk (Figure 6A and Table S6).

**Figure 6.**
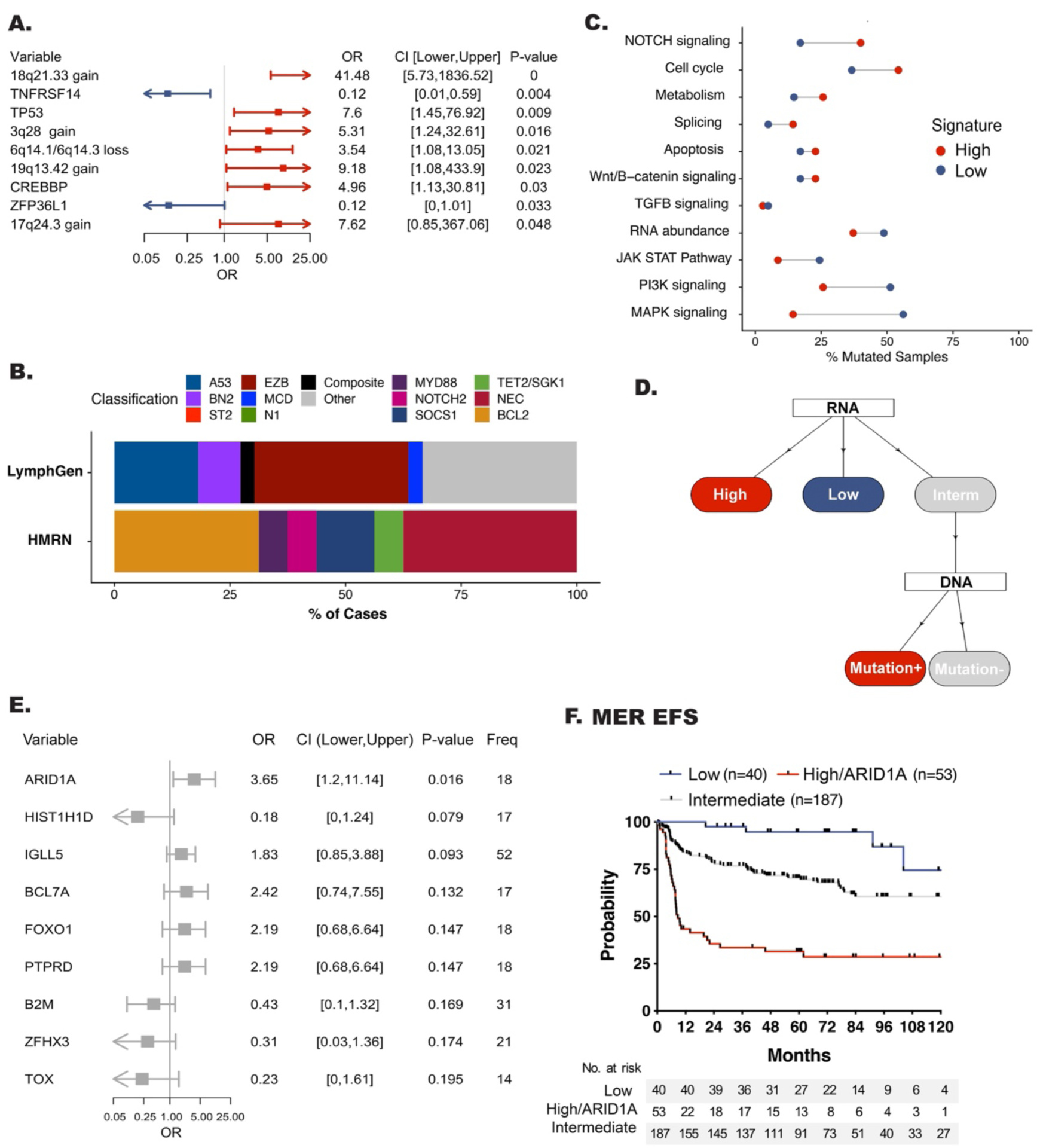
Genetic Features of High Risk DLBCL. A. Forest plot showing enrichment of mutations and copy number events between the high and low risk groups. B. Dot plot showing the percentage of samples which have mutations in the represented pathway. Red dots represent the percentage of samples in the high risk group while blue dots represent the percentage of cases in the low risk group. Shown pathways have at least a 1.3 fold increase or decrease between both groups. C. Schematic representation of decision tree approach for identification of EFS24 mutations in intermediate risk DLBCL. D. Forest plot showing enrichment of mutations in the intermediate group, comparing EFS24 fail vs EFS24 achieve. E. Kaplan Meir curve showing event free survival of high risk DLBCL with the inclusion of *ARID1A* mutations.

Classification of by LymphGen and HMRN (Figure 6B and Figure S10C) revealed that the high risk cases are spread across the molecular classifiers. Because DLBCL is genetically heterogeneous and not dominated by single mutations, it is possible to miss the importance of less frequent variants that may play a role in aggressive biology. We therefore performed a pathway analysis using all genes previously reported to be mutated in lymphoma (n=268) as well as the PanCancer gene list (n=184) from maftools (Table S7).^11, 12, 18, 32, 33^ This analysis revealed an enrichment of mutations in genes related to Notch signaling, the cell cycle, splicing, and metabolism pathways, when compared to low risk (Figure 6C). Conversely, there was enrichment of mutations in genes related PI3-kinase, Jak/Stat, and MAP-kinase in the low risk cases.

In our ndDLBCL cohort the high risk signature captured 36% of EFS24 failures, but only 5% of EFS24 achievers. However, a significant number of EFS24 failures remain in the intermediate risk group. Therefore, we wanted to determine if we could integrate our genetic data with our risk signatures to further refine our ability to capture additional cases (Figure 6D). We first ran a mutation enrichment analysis on the intermediate risk group to identify mutations associated with EFS24 failure and identified that *ARID1A* (Figure 6E). Next, we categorized patients who had both RNA-seq and WES data as low, intermediate, or high risk and/or *ARID1A* mutation. Similar to prior analysis, compared to low risk the new integrated high risk signature was associated with EFS (HR, 13.49 [95% CI 4.97-37.96] *P* < .001) (Figure 6F.) These findings were validated in independent data sets from BCCA (PFS, HR 7.3, [95% CI 1.67-32.00] *P*=.008), and Duke (OS, HR 2.93, [95% CI 1.55-5.56] *P =* .001), and REMoDL-B (PFS, HR 2.59, [95% CI 1.29-5.17] *P* =.007) (Figure S11 A-C). Overall, the integrated high risk signature captured 45% of EFS24 failure in the MER cohort, 34% in BCCA, and 37% in REMoDL-B, an improvement compared the RNA signature alone (Figure S11D).

## DISCUSSION

In recent years, several molecular and immune classification systems have been developed to subgroup DLBCL, which has advanced the understanding of biological pathways driving this disease. However, our results suggest that these known clusters fail to identify patients with a early clinical failure, limiting their use in clinical decision making. Using a multiomic approach on a highly annotated cohort of ndDLBCL, we now describe a risk signature that simultaneously captures risk status and aggressive biology defined by metabolic and TME depleted expression profiles. In a simultaneous analysis of this data, we performed an unsupervised analysis on transcriptomics features from ndDLBCL patients and identified 7 clusters, one called A7 (Aggressive lymphoma 7) with poor prognosis and defined by ABC COO and high myc expression (currently under review). Interestingly, our high risk gene expression signature shared overlap with only 17% of A7 cases, suggesting that unique aggressive features were detected by each approach.

Our finding that high risk DLBCL tumors were driven by a metabolic signature are supported by prior gene expression studies where Monti et al ^34^ identified a subgroup of DLBCL, OxPHOS-DLBCL, defined by a dysregulation if genes belonging to the mitochondrial oxidative phosphorylation pathway (OxPhos)^34^ with further analysis suggesting that those tumors develop an independent nutrition mechanism.^35^ However, the association of OxPHOS-DLBCL with outcome has not been fully explored. The metabolic shift being detected by our high risk signature may be consistent with the “Warburg Effect” ^36^ a well know mechanism of cancer progression that is often associated with aggressive disease. Our high risk cases also displayed a depleted TME signature ^37^ suggesting poor infiltration of tumor specific T cells. There is a growing literature suggesting that a lack of TME involvement has a negative impact on outcome,^16, 17^ Kotlov et al, reported that the highest number of non-responders on standard chemotherapy where classified as TME depleted.^17^ Our high risk signature is unique in that it simultaneously captures both metabolic and TME dysregulation allowing for early capture of aggressive DLBCL with potentially heterogenous biologic programs.

Several studies have attempted to identify the prognostic value of single genetic alterations,^38, 39^ yet there is little consensus between studies. However, our data do align with previous findings on *TP53* alterations, which have been shown to be prognostic of inferior survival in DLBCL.^39–42^ Although, *TP53* mutations alone were not prognostic in our cohort, likely due to the fact that not all *TP53* mutations have the same biologic impact. Because *TP53* mutations are well known to be associated with metabolic rewiring and chemoresitance, we hypothesize it may play an important role in driving the metabolic signature identified in our study.^42–46^ Beyond *TP53*, our analysis highlights a role for *ARID1A* in aggressive disease biology. *ARID1A* mutations are associated with both tumor suppression and tumor initiation in many malignancies, including DLBCL.^47, 48^ In addition to being an important chromatin modifier, *ARID1A* is involved in double strand break repair, homologous recombination, and mismatch repair pathways.^49–53^ *ARID1A* can also directly bind *TP53* to enhance its activity^54^, thus loss of *ARID1A* may act like a tumor suppressor and have a negative prognostic impact even in the absence of *TP53* alterations.

Moving forward, our signature and proposed classification approach (Figure S11E) may have important clinical implications. While not a primary focus of this manuscript, our risk classifier identified cases with a low risk of having an early event. This may be a subgroup of patients that will benefit from standard of care treatment with R-CHOP and may be spared from use of more toxic or expensive therapies.

Identification of cases at diagnosis with our high risk signature could select patients appropriate for clinical trials of earlier use of CAR-T; earlier identification could allow for sooner CAR-T manufacturing reducing the percentage of patients with progression prior to receiving CAR-T, a key barrier to this therapy. We also identified several important biological pathways that may be directly targetable. Pre-clinical studies have shown that cell lines with *ARID1A* mutations are sensitive to EZH2 inhibitors^57^, such as tazemetostat, which are currently approved for the treatment of FL and are in clinical trial development for DLBCL.

In summary, to our knowledge, this is the first classification system to use novel and integrative computational approaches to identify a multiomic signature of early clinical failure. Our signature captures important clinical and pathological characteristics, individual molecular alterations, and biological pathways in one signature for patient stratification and clinical management.

## Data Availability

All data produced in the present study are in the process of being made available for public access

## ACKOWLEDGEMENTS

This work was supported in part by the NIH/NCI grants SPORE-P50 CA97274 (J.R. Cerhan and A.J.Novak), R01 CA212162-01A1 (A.J. Novak and J.R. Cerhan), U01 CA195568 (J.R. Cerhan), T32AI007425 (J.E. Krull) and T32AI170478 (M.A. Hopper).

## DATA SHARING STATEMENT

RNA and DNA sequencing data is in the process of being submitted to EGA.

## AUTHOR CONTRIBUTIONS

### Conception and design

Kerstin Wenzl, Matt Stokes, James R. Cerhan, Anita K. Gandhi and Anne J. Novak

### Financial support

James R. Cerhan, Anita K. Gandhi and Anne J. Novak

### Provision of study materials or patients

Lisa Rimsza, Brian K. Link, Stephen M Ansell, Thomas M. Habermann, Andrew L. Feldman, Rebecca L. King, Grzegorz Nowakowski, James R. Cerhan, Anita K. Gandhi and Anne J. Novak

### Collection and assembly of data

Kerstin Wenzl, Matt Stokes, Joseph P. Novak, Vivek Sarangi, Raphael Mwangi, Maria Ortiz, Nicholas Stong, C. Chris Huang, Matthew J. Maurer

### Data analysis and interpretation

Kerstin Wenzl, Matt Stokes, Joseph P. Novak, Allison M. Bock, Sana Khan, Melissa A. Hopper, Jordan E. Krull, Abigail R. Dropik, Janek S. Walker, Vivek Sarangi, Raphael Mwangi, Maria Ortiz, Nicholas Stong, C. Chris Huang, Matthew J. Maurer, Susan L. Slager, Yan Asmann, Patrizia Mondello, Ryan Morin, James R. Cerhan, Anita K. Gandhi and Anne J. Novak

### Manuscript writing

All authors

### Final approval of manuscript

All authors

### Accountable for all aspects of the work

All authors

## Notes

### Competing Interest Statement

Anne Novak has received research funding from Bristol Myers Squibb

### Author Declarations

This study was approved by the Mayo Clinic IRB and patients provided written consent at study enrollment or or had waiver of consent for use of clinical samples.

