## Supplementary material for "Multiomic Analysis Identifies a High-Risk Metabolic and TME Depleted Signature that Predicts Early Clinical Failure in DLBCL": Data Supplement Figures

### I. Supplemental Methods

#### Study Populations

All samples had a diagnosis of DLBCL. All FFPE sample used for DNA and RNA extraction were reviewed by a Mayo Clinic hematopathologist prior to sectioning. Cell of origin (COO) was determined on available samples by the Lymph2Cx assay (NanoString, n=326)[1] or from RNA-Seq data (n=86) using the method described by Reddy et al. *MYC* (n=318), *BCL2* (n=317), and *BCL6* (n=313) FISH was performed on available samples as previously described.[2,3] Double Hit signature status (DH-sig) was determined according to Ennishi et al.[4]

#### DNA Sequencing and Analysis

Tumor DNA was extracted from formalin-fixed, paraffin embedded (FFPE) tissue sections and whole exome sequencing (WES) of all samples was performed at Expression Analysis using the Agilent SureSelect XT AllExon v5 + UTR kit and sequencing was carried out on an Illumina NovaSeq, 100 x 2 paired end reads. GATK best practices workflow was followed using Sentieon (v201808.05) implementations of picard and BWA. Reads were trimmed with cutadapt and then aligned to human genome reference build 38 using BWA mem (0.7.17). For calling single-nucleotide variants (SNVs) and INDELs GATK v4.0.12 mutect2 was used. Post alignment and somatic mutation calling, common variants which with a frequency higher than 10% percent in ExAC or gnomAD were removed. Mutations included required a depth of at least 10 in both tumor and normal, greater than 5% allele frequency in the tumor, less than 5% in the normal, and a minimum alternate allele depth of 3. For copy number analysis, OncoScan (n=213) or WES (n=174) files were used for the copy analysis; comparison of copy number calls across the two platform has been previously published.[5] Bam files for tumor and germline sample and OSCHP files were loaded into the software and aligned to human genome reference build 37 (GRCh37).

For HMRN classification, mutation, amplification, and deletion data were modeled as a finite mixture of Bernoulli distributions using R code provided by the authors ( https://github.com/ecsg-uoy/DLBCLGenomicSubtyping) 360 cases met submission criteria for HMRN.[6] For LymphGen classification, data were prepared and submitted for classification as instructed by the online tool (https://llmpp.nih.gov/lymphgen/index.php).[15] 369 cases met submission criteria for LymphGen. Oncoprints for mutation and copy number data were generated using maftools and complex heatmaps.[7,8]
Mutations in the oncoprint are grouped into Missense (Missense mutation), Truncating (Frame Shift Del, Frame Shift Ins, Splice Site, Translation Start Site, Nonsense Mutation, Nonstop Mutation), In-Frame (In Frame Del, In Frame Ins) and Multi-hit mutations. Oncogenic signaling pathway analysis was carried out using maftools.[7]

#### RNA Sequencing and Analysis

RNA sequencing was performed using the Illumina TruSeq RNA Exome Kit (Illumina) for library preparation, sequencing platform HiSeq 4000, 100 x 2 paired end reads. The RNA sequencing paired-end reads fastq files were processed as previously described (Stokes et al.). Briefly, the sequencing data were processed on a cloud-based platform at Bristol Myers Squibb (BMS). Fastq files were aligned to the human genome reference build 38 (GRCh38) using the Star aligner method.[9] Quantification of the aligned RNA sequencing data was carried out using salmon.[10]

Weighted Gene Correlation Network Analysis (WGCNA) was performed using all protein coding genes, with the exclusion of X, Y and M chromosomes for the network analysis.[11] To calculate the similarity matrix between genes across all samples, Pearson's correlation was used. To achieve scale-free topology the parameter (β) was set to 17. The similarity matrix was transformed to an adjacency matrix, then the topological overlap matrix (TOM) and the dissimilarity topological overlap matrix (1-TOM) were computed. In a final step, hierarchical clustering and dynamic tree cut were used to reveal the co expression modules. The minimum model size for our data set was set to 15 genes and the cut size was 0.25. The module (specifically, module eigengene, which is the first principal component of the module expression) was correlated with clinical traits was done using Pearson correlation. For visualization of the greenyellow module correlation network the R tool igraph with the Fruchtman Reingold layout was used.[12]

The count data were used to carry out differential expression analysis (DEG) analysis using the edgeR R package.[13] Only genes which had an of FDR < 0.05 were considered significant and were used for further analysis. The cystoscape module GluGo was used for pathway analysis of the WGCNA results and the R package pathfindR was used for pathway analysis of the differential gene expression analysis using KEGG pathway and Gene Ontology (GO) annotation.[14-16] Overrepresentation analysis of the RNA signature was done using the gprofiler2 R tool.[17] TME26 was scored according to Risueño et al.[18] For immune deconvolution, CIBERSORTx was run on the on log2(TPM+1) gene expression values.[19] The LM22 signature matrix was used and the data were permuted 500 times. The final data were reported as absolute abundances for each cell type. We also analyzed our data using the Lymphoma Microenvironment (LME) tool (https://github.com/BostonGene/LME) and EcoTyper (https://ecotyper.stanford.edu/lymphoma/).[20,21]

**RNA Risk Signature Scoring and Validation**

The R tool singscore was used to score our individual cases for the signature and each patient was assigned a totalscore based on expression of both up- and down-regulated genes.[22] The z-score for the totalscore was calculated and cases were classified as low, intermediate, or high risk based on +/- one standard deviation away from the mean. Three validation cohorts for the RNA signature were used. For the BCCA cohort the input matrix included all genes (n = 54,397) and 384 RNA signature genes. The Duke input matrix included 14,513 genes and 387 RNA signature genes. The input matrix for the REMoDL-B dataset included 12,736 and 335 RNA signature genes.

### II. Supplemental Figures

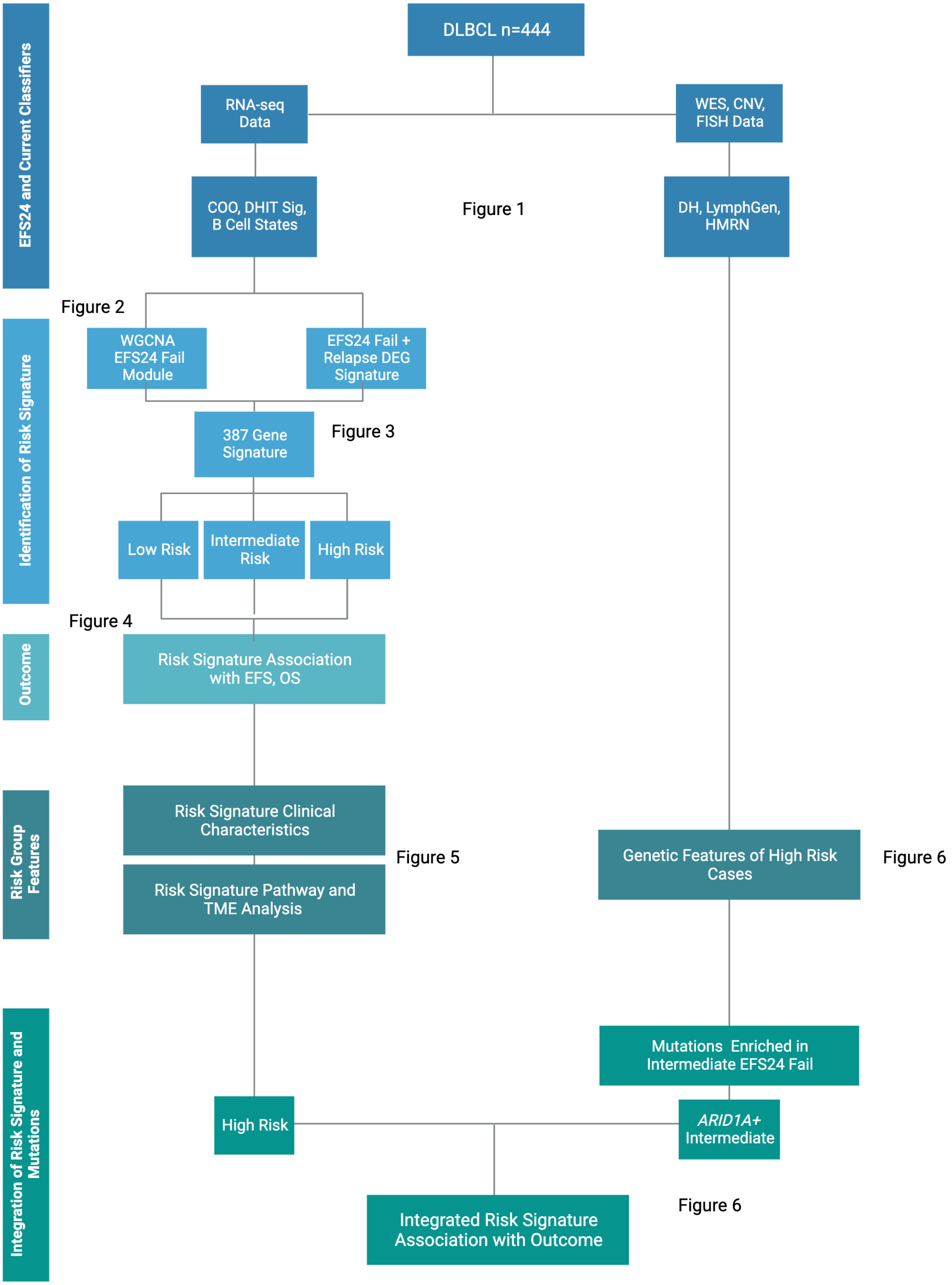


**Supplemental Figure 1. Study Design.** The overall schematic of how the study was performed is shown. Created with BioRender.com.

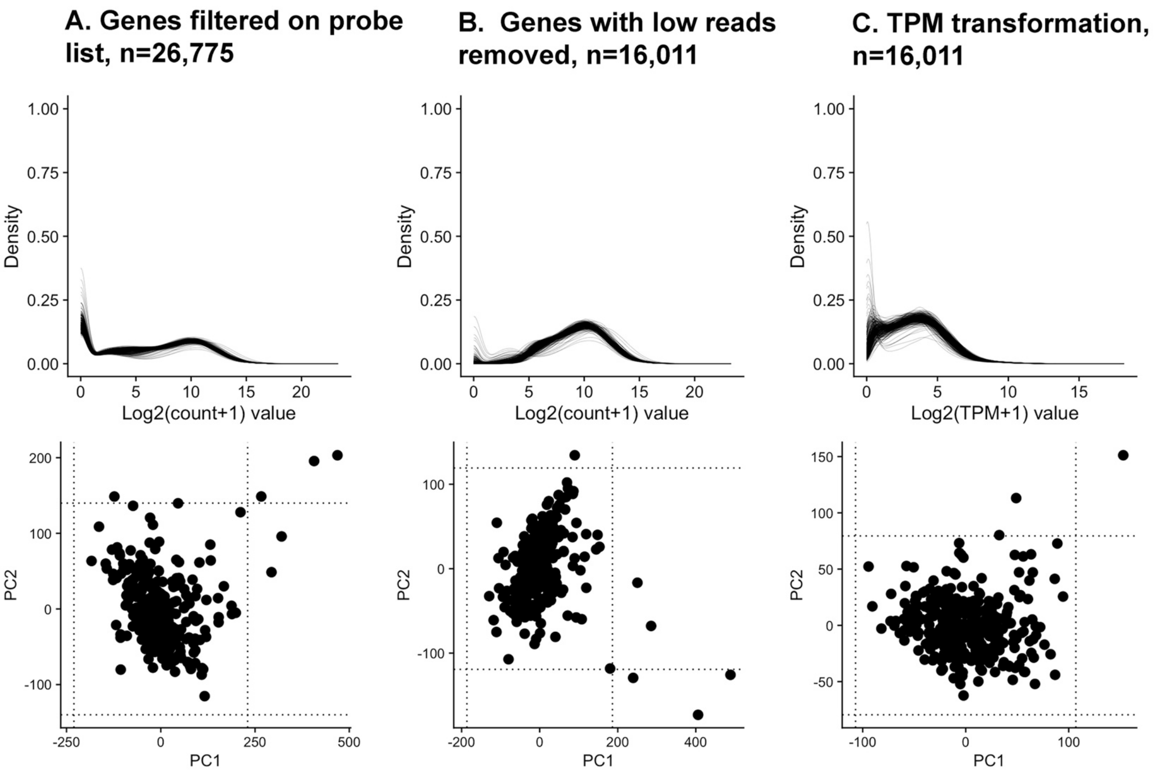


**Supplemental Figure 2. Quality control of RNA sequencing data.** Density (upper panel) and PCA (lower panel) plots from 323 ndDLBCL samples showing A. Genes included on probe list for the Illumina TruSeq RNA Exome Kit. B. Genes with a median read count less than were removed, and C. Distribution of samples after TPM transformation. This analysis resulted in the exclusion of 2 samples for a final cohort size of 321 cases.

**A.**

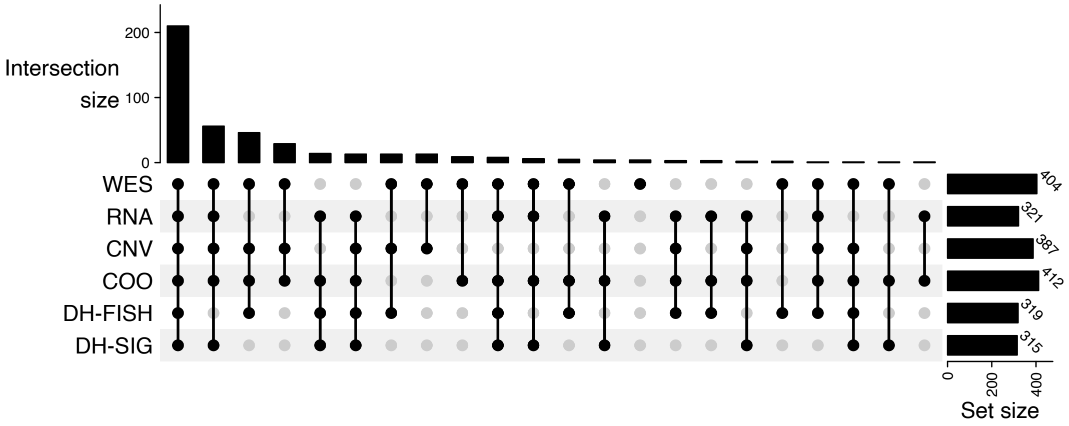


**B.**

| Patient Characteristics | Total (N=444) |
|---|---|
| **Age at Diagnosis** | |
| Mean (SD) | 62.9 (13.69) |
| Median | 64.5 |
| IQR | 55.0, 72.5 |
| Range | 18.0, 93.0 |
| **Age Group**, n (%) | |
| <=60 | 166 (37.4%) |
| >60 | 278 (62.6%) |
| **Gender**, n (%) | |
| Male | 251 (56.5%) |
| Female | 193 (43.5%) |
| **PS Group**, n (%) | |
| <2 | 372 (84.2%) |
| >=2 | 70 (15.8%) |
| Missing | 2 |
| **Ann Arbor Stage**, n (%) | |
| I-II | 178 (40.2%) |
| III-IV | 265 (59.8%) |
| Missing | 1 |
| **Extranodal Sites**, n (%) | |
| 0-1 extranodal sites | 351 (79.1%) |
| 2 or more extranodal sites | 93 (20.9%) |
| **LDH Group**, n (%) | |
| Elevated | 214 (51.3%) |
| Not elevated | 203 (48.7%) |
| Missing | 27 |
| **IPI**, n (%) | |
| 0 | 48 (10.8%) |
| 1 | 108 (24.3%) |
| 2 | 121 (27.3%) |
| 3 | 109 (24.5%) |
| 4 | 49 (11.0%) |
| 5 | 9 (2.0%) |

| Patient Characteristics | Total (N=444) |
|---|---|
| **IC Treatment Group**, n (%) | |
| R-CHOP | 306 (68.9%) |
| R2-CHOP | 53 (11.9%) |
| MR-CHOP | 31 (7.0%) |
| R-EPOCH | 23 (5.2%) |
| Other IC | 14 (3.2%) |
| ER-CHOP | 10 (2.3%) |
| RAD-RCHOP | 4 (0.9%) |
| RCHOP/Zevalin | 3 (0.7%) |
| **Cell of Origin**, n (%) | |
| ABC | 131 (31.8%) |
| GCB | 232 (56.3%) |
| Unclassified | 49 (11.9%) |
| Missing | 32 |
| **EFS Status**, n (%) | |
| Event | 168 (37.8%) |
| No Event | 276 (62.2%) |
| **EFS24 Status**, n (%) | |
| Achieved EFS24 | 332 (74.8%) |
| Failed to Achieve EFS24 | 112 (25.2%) |
| **Alive Pts Time to Follow-Up (Mo)** | |
| Mean (SD) | 91.4 (42.33) |
| Median | 83.8 |
| IQR | 59.4, 119.2 |
| Range | 0.2, 195.9 |

**Treatment Definitions**

| | |
|---|---|
| R-CHOP | Rituxan, Cyclophosphamide, Doxorubicin, Vincristine, Prednisone |
| R2-CHOP | R-CHOP + Lenalidomide |
| MR-CHOP | R-CHOP + High-dose Methotrexate |
| R-EPOCH | R-CHOP + Etoposide Phosphate |
| ER-CHOP | R-CHOP + Epratuzumab |
| RAD-RCHOP | R-CHOP + Radiation |
| RCHOP/Zevalin | R-CHOP + Zevalin |

**Supplemental Figure 3. Study Cohort Data Availability and Patient Characteristics.**

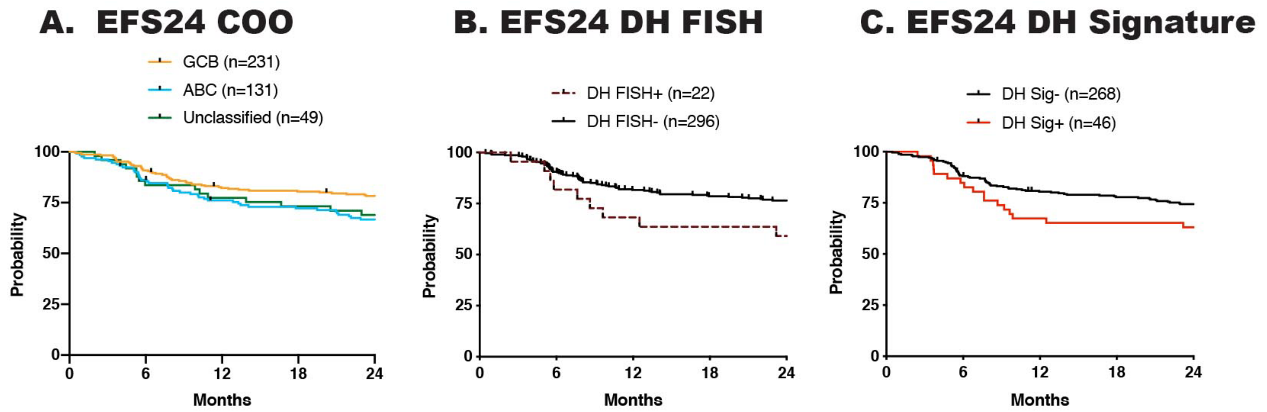


**Supplemental Figure 4. Association of EFS24 with COO, DH-FISH, and DH-Signature Status**. EFS24 survival curves for ndDLBCL cases according to their A. COO, B. DH-FISH, and C. DH-Signature status.

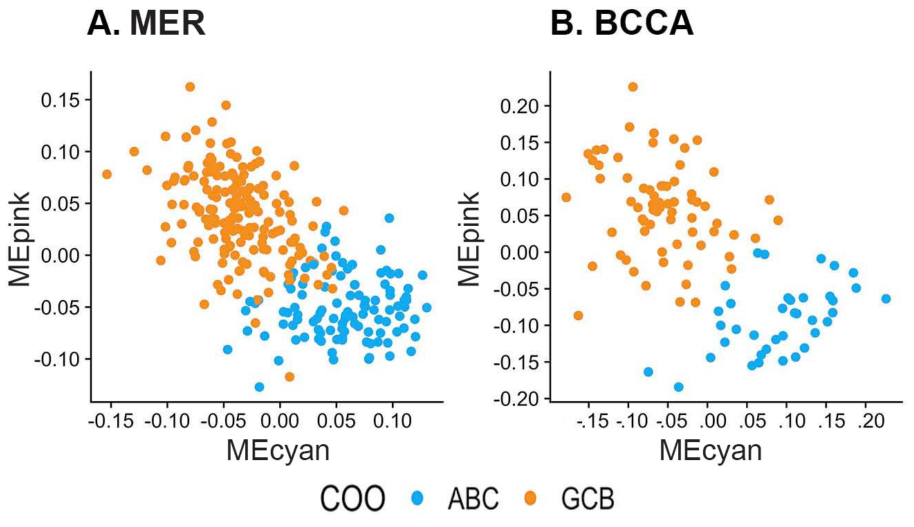


**Supplemental Figure 5 WGCNA Eigengene Values Correlate With COO Calls.** Scatterplots of eigengene values for the pink (correlated with GCB) and cyan (correlated with ABC) WGCNA modules in the A. Mayo (n=279) and B. BCCA (n=107) cohorts. Blue dots indicate an ABC COO and orange dots indicate a GCB COO determined by the Lymph2Cx assay or the Reddy et al method. Higher eigengene values for the pink module associated with the GCB and higher eigengene value for the cyan module associated with ABC in both cohorts.

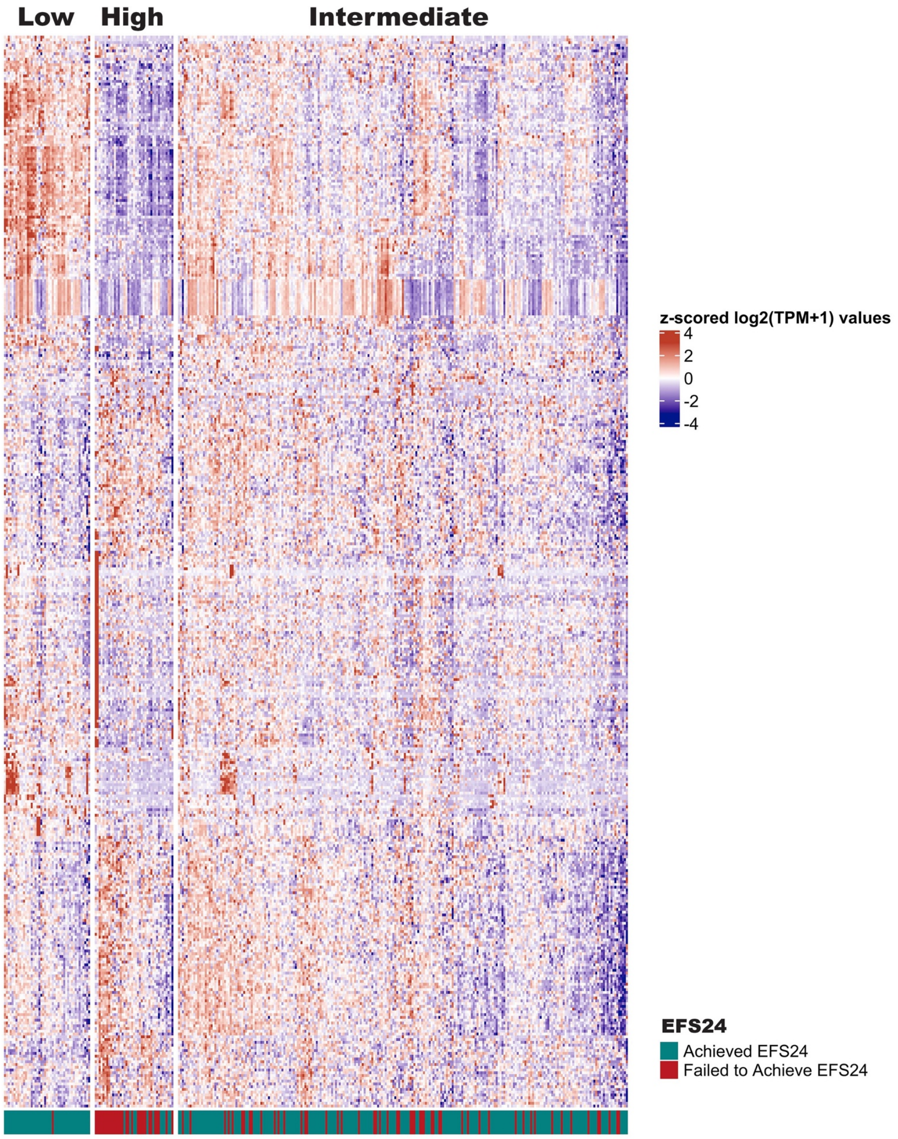


**Supplemental Figure 6. Heatmap of RNA Risk Signature Genes.** Heatmap of gene expression for the 387 genes used for classification of samples (n=321) into high, low or intermediate risk.

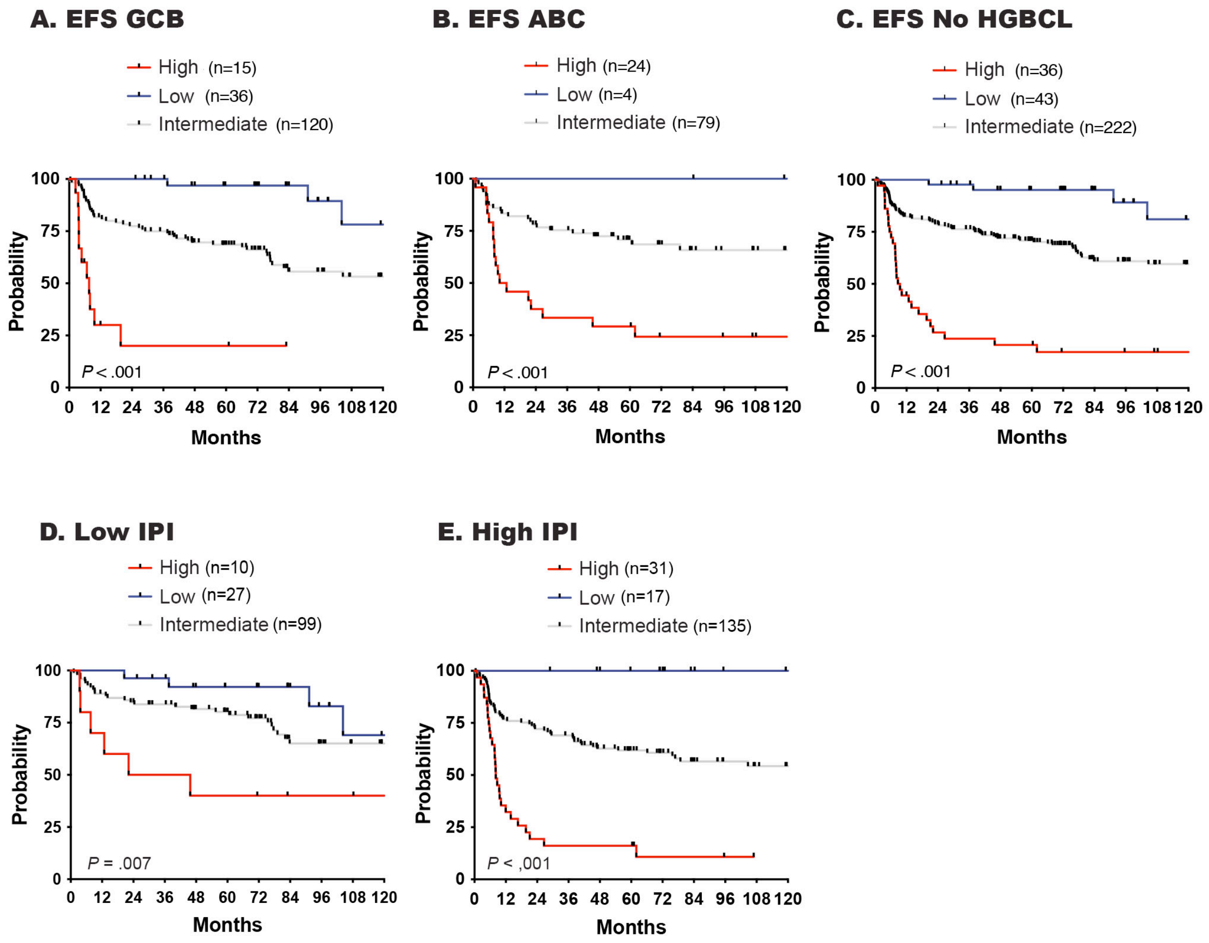


**Supplemental Figure 7. Kaplan Meier Analysis of RNA Risk Signature Scored Patients**. EFS survival curve of high, low and intermediate risk patients in A. GCB cases, B. ABC cases, C. HGBCL cases excluded, D. Low IPI cases, and E. High IPI cases. Log-rank test was used to calculate the *P*-value.

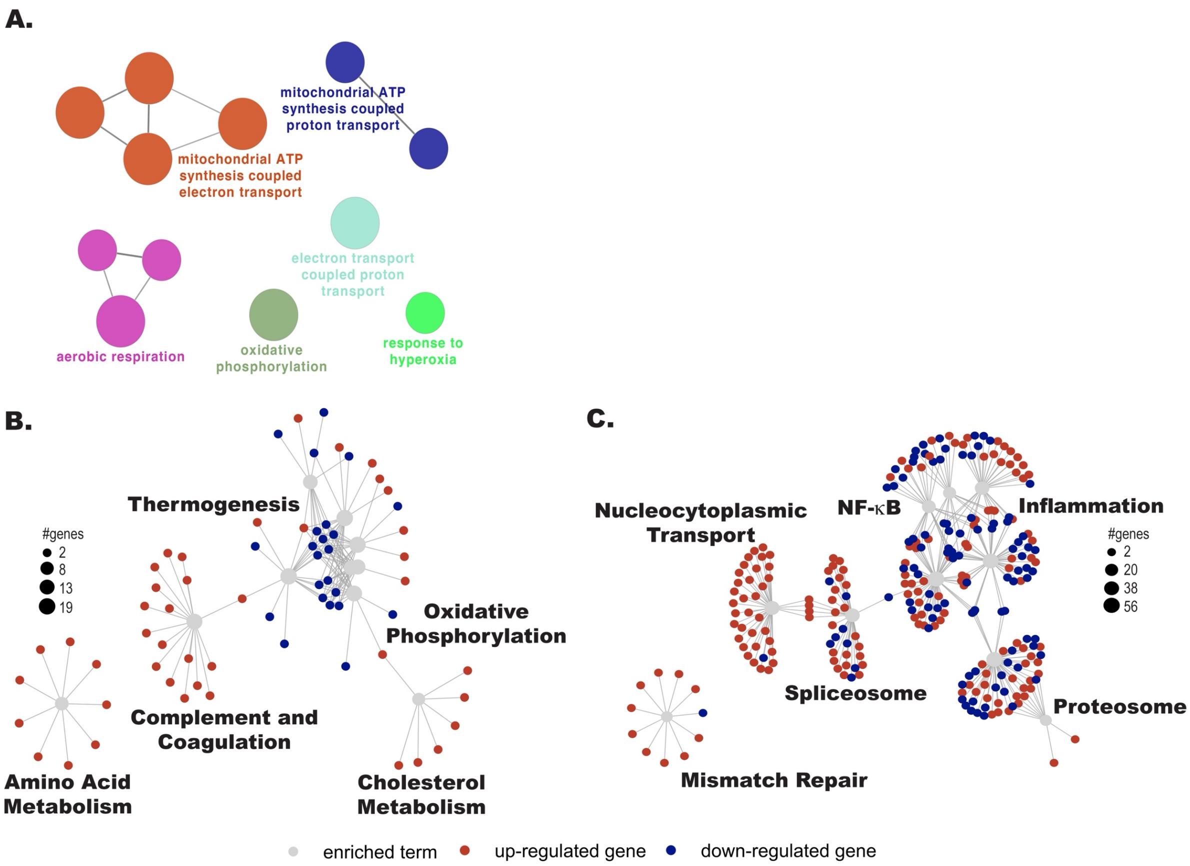


**Supplemental Figure 8. Pathway Analysis of WGCNA and DEG Analysis.** A. Overrepresentation analysis of genes from the greenyellow WGCNA module performed using the Cytoscape module GluGo. B. Pathway analysis was performed on the differentially expressed genes between B. EFS24 achieved and failed, and C. EFS24 achieved and rrDLBCL using PathfindR. Top 10 significant pathways (*P* < .05) are shown.

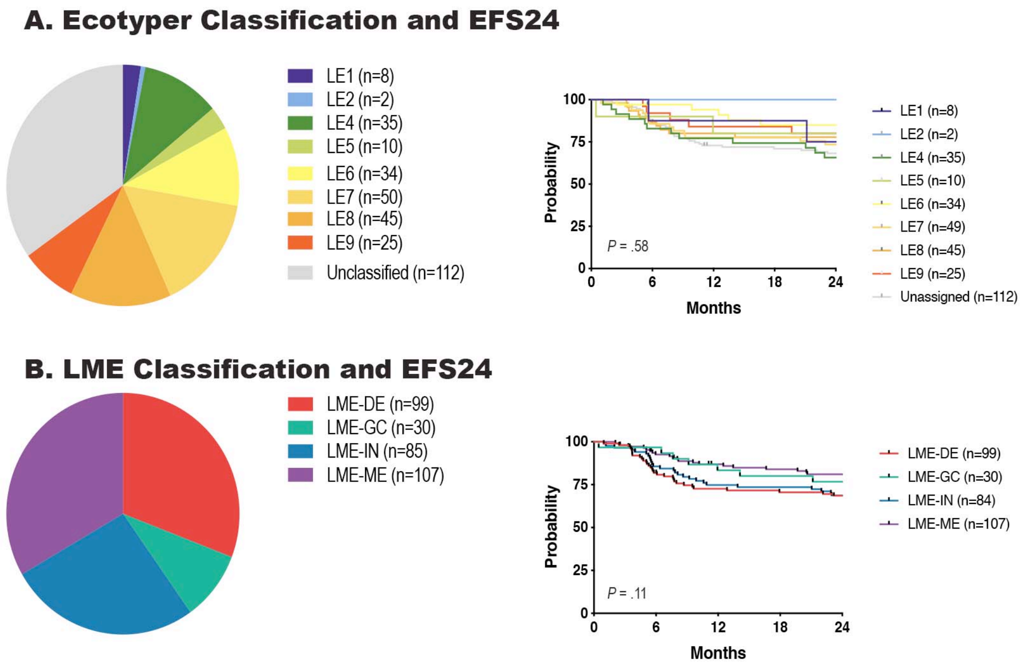


**Supplemental Figure 9 Lymphoma Ecotype and LME Classification and Outcomes in Mayo ndDLBCL.** A. Pie chart showing distribution and EFS24 outcome of cases according to Ecotyper classification. B. Pie chart showing distribution and EFS24 outcome according to LME classification Log-rank test was used to calculate the *P*-value.

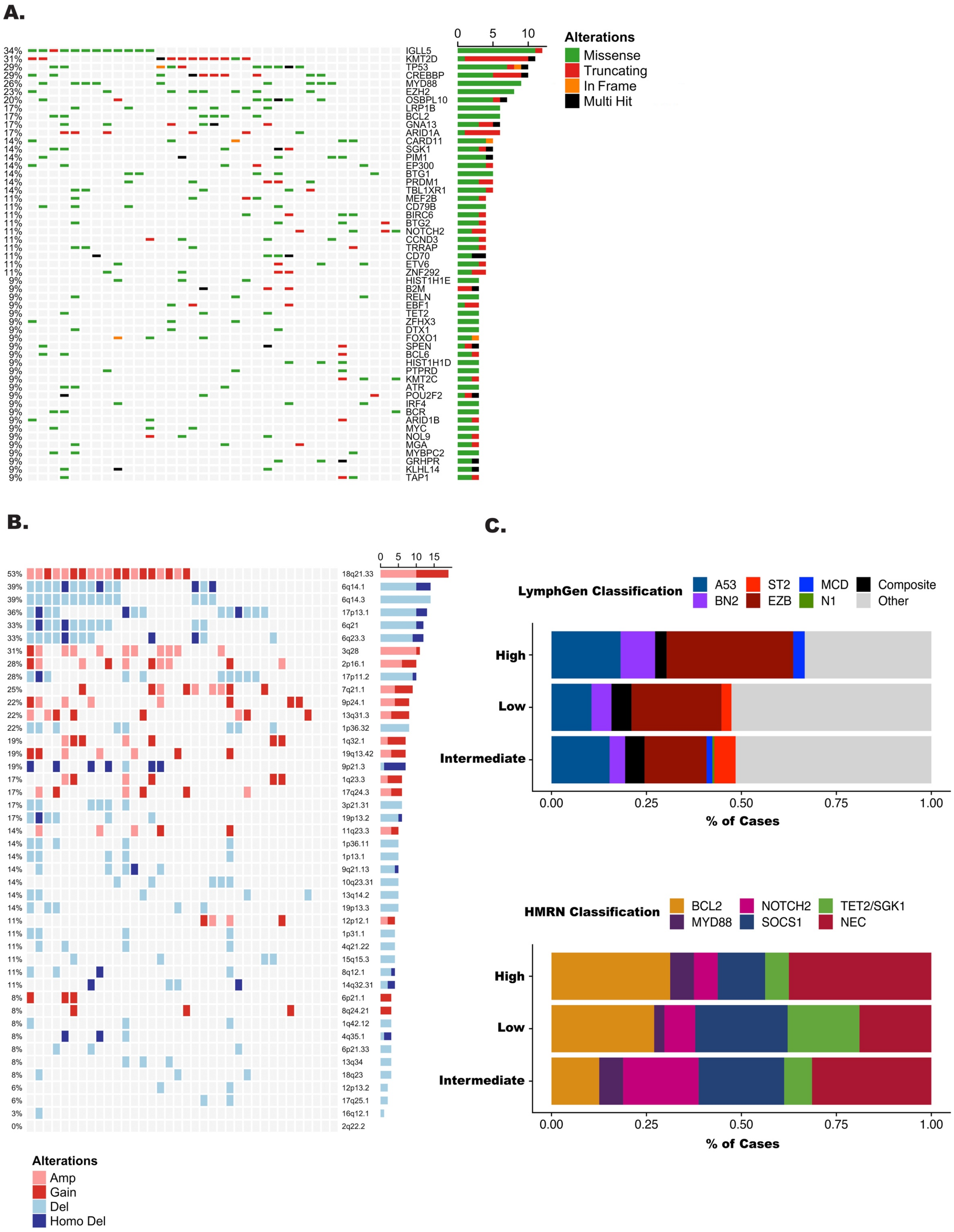


**Supplemental Figure 10 Genomic Landscape of High Risk DLBCL**. A. Oncoprint of lymphoma driver genes in the high risk group using a 9% frequency cutoff. B. Oncoprint of copy number events in the high risk group. LymphGen (top panel) and HMRN (bottom panel) classification of cases in the high, low, and intermediate risk group. C. LymphGen and HMRN classification of high, low, and intermediate risk groups.

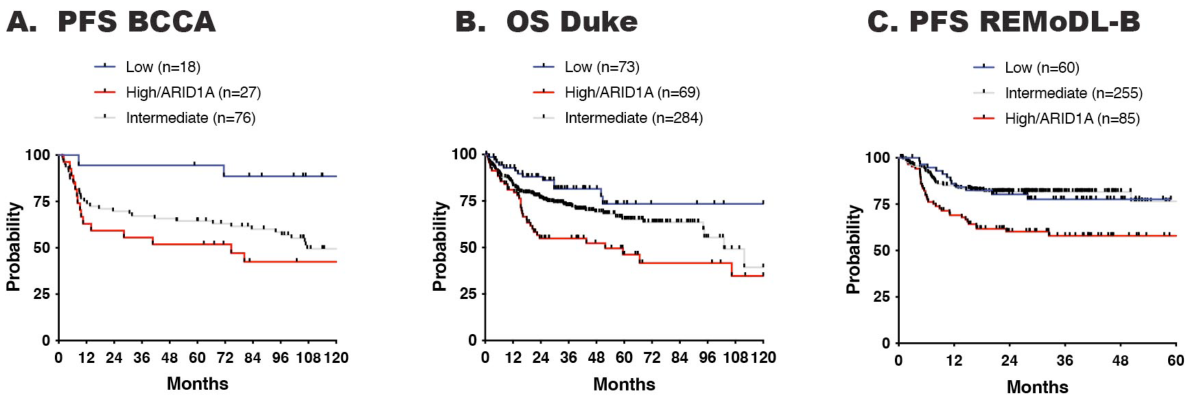


**D.**

| | MER | BCCA | REMoDL-B |
|---|---|---|---|
| High Risk Signature | 36% | 25% | 25% |
| High Risk Signature and/or *ARID1A+* Intermediate | 45% | 34% | 37% |

**E.**

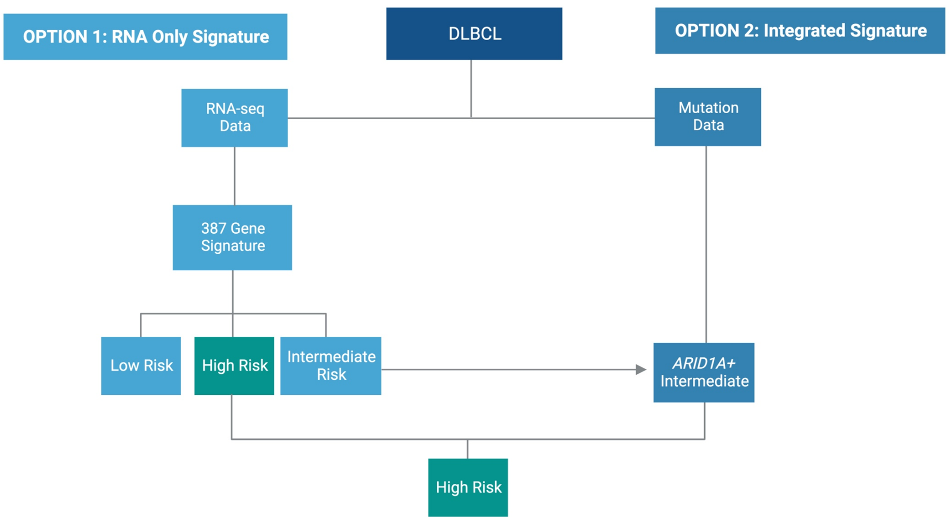


**Supplemental Figure 11 Survival Curves of Validation Cohorts Classified by Risk Signature and *ARID1A* Mutations**. A. Progression free survival curves for the BCCA cohort. B. OS survival curves for the Duke cohort. C. Progression free survival curves for the REMoDL-B Rituxan arm cohort (WES data available). D. Percent of cases that had an event before 24 months in the MER, BCCA, and REMoDL-B cohorts identified by the RNA high risk signature alone or the integrated high risk signature including *ARID1A* mutations. E.  Proposed approach to identifying high risk cases. Created with BioRender.com.
